# Profiles of cognitive change in preclinical Alzheimer’s disease using change-point analysis

**DOI:** 10.1101/19009696

**Authors:** Owen A Williams, Yang An, Nicole M Armstrong, Melissa Kitner-Triolo, Luigi Ferrucci, Susan M Resnick

**Affiliations:** Laboratory of Behavioral Neuroscience, National Institute on Aging, Baltimore, Maryland 21224, USA; Longitudinal Studies Section, Translational Gerontology Branch, National Institute on Aging, Baltimore, MD, USA

**Keywords:** Preclinical, Cognitive Decline, Change-point analysis, Visuospatial ability, Alzheimer’s Disease

## Abstract

**Introduction:** Change-point analyses are increasingly used to identify the temporal stages of accelerated cognitive decline in the preclinical stages of Alzheimer’s Disease (AD). However, statistical comparisons of change-points between specific cognitive measures have not been reported.

**Methods:** 165 older adults (baseline age range: 61.1-91.2) from the Baltimore Longitudinal Study of Aging developed AD during follow-up. Linear and non-linear mixed models were fit for 11 cognitive measures to determine change-points in rates of decline before AD diagnosis. Bootstrapping was used to compare the timing of change-points across cognitive measures.

**Results:** Change-points followed by accelerated decline ranged from 15.5 years (Card Rotations) to 1.9 years (Trail-Making A) before AD diagnosis. Accelerated decline in Card Rotations occurred significantly earlier than all other measures, including learning and memory measures.

**Discussion:** Results suggest that visuospatial ability, as assessed by Card Rotations, may have the greatest utility as an early predictive tool in identifying preclinical AD.

## Introduction

Alzheimer’s disease (AD) is a progressive disease with a long preclinical phase in which pathological markers are present for years and even decades before clinical symptoms [1]. Decline in episodic memory is a hallmark of AD, but other cognitive domains are also vulnerable to AD. [2] Understanding the temporal stages of the early acceleration of declines in various cognitive domains in preclinical AD is important for identifying individuals vulnerable towards accumulating AD pathology and for characterizing AD progression prior to symptom onset.

The early preclinical phase of AD is characterized by beta amyloid (Aβ) and phosphorylated tau accumulation with subsequent acceleration of brain atrophy in the absence of clinical symptoms. [1, 3-5] With multiple anti-amyloid clinical trials failing to show that removal of Aβ is associated with improved cognitive outcomes, [6] one argument is that patients are not being targeted early enough. [6, 7] The failures of clinical trials at later disease stages have led to increasing focus on the earlier phases of disease, including the asymptomatic preclinical stage, with the hope that treatments at this stage may be more effective. [8, 9] Thus, it is critical to define the earliest, and possibly subtle, changes in cognitive performance in preclinical AD to identify individuals who would have the greatest potential to benefit from clinical interventions. To fully characterize cognitive changes in preclinical AD, it is important to examine a broad range of cognitive domains and neuropsychological measures that may be sensitive to the earliest changes.

One way of investigating the timing of cognitive decline prior to clinical AD diagnosis is to use change-point analyses. Changepoint methods align participants by anchoring them at time of diagnosis to then examine trajectories of variables of interest retrospectively for timepoints of change prior to clinical diagnosis. Change-points are identified using piece-wise linear components separated by knots delineating between intervals with differing rates of change. [10, 11] Previous studies using change-point analyses in AD focused on verbal memory, [12, 13] reporting steeper declines in Immediate Recall, measured by the picture version of the Free and Cued Selective Reminding test, between 1 and 8.1 years before clinical diagnosis. However, memory is not the only cognitive domain subject to decline prior to AD onset [2] and other domains have shown early change-points. In a systematic review of change-point studies in dementia and AD, [14] the measure with the earliest change-point, at 9.6 years before AD diagnosis, was the Block design test assessing visuospatial ability. [15] Measures associated with other domains, i.e., language fluency and executive function showed change-points detected at 6.8 years [16] and 2.9 years [13] prior to AD diagnosis respectively.

While the systematic review by Karr et al. [14] allows for a cursory comparisons of change-points between measures associated with various cognitive domains, the authors highlight various methodological differences between studies that make it difficult to draw conclusions from such comparisons. For example, the maximum length of longitudinal testing prior to AD diagnosis ranged from 9-30 years with frequency of visits varying between studies. Furthermore, the mean baseline ages in all studies ranged from 70-82 years, and analyses were adjusted by different sets of covariates. Therefore, the temporal sequence of changes in different cognitive domains in preclinical AD remains unclear.

To elucidate the temporal sequence of cognitive changes prior to clinical AD onset, we investigated changes in rates of decline on multiple cognitive measures, representing specific cognitive domains, in individuals who eventually developed AD. The aims of this study were to identify how many change-points best characterize the trajectories of change in performance on several cognitive measures and to find the earliest point in time before AD diagnosis that accelerated decline in performance could first be detected for each measure.

## Methods

### Participants

Participants were from the Baltimore Longitudinal Study of Aging (BLSA), a longitudinal study started in 1958. [17] Participants were community-dwelling volunteers who were healthy at enrollment. During each visit, they received comprehensive psychological evaluations. For this study, we selected participants who were diagnosed with AD during follow-up and only used data from visits when they had complete neuropsychological testing data across the 11 measures investigated.

### Clinical diagnosis of Alzheimer’s disease

Clinical and neuropsychological data from each participant were reviewed at a consensus case conference if their clinical dementia rating score [18] was 0.5 or greater or if they had more than three errors on the Blessed Information-Memory-Concentration Test, [19] and participants were evaluated by case conference upon death or withdrawal. MCI status was determined using the Petersen criteria. [20] Diagnoses of dementia and AD were based on criteria outlined in the Diagnostic and Statistical Manual of Mental Disorders, third edition, revised [21] and the National Institute of Neurological and Communication Disorders and Stroke – Alzheimer’s Disease and Related Disorders, [22] respectively.

### Neuropsychological measures

Participants were administered a comprehensive battery of cognitive measures assessing verbal learning and memory, figural memory, attention and processing speed, executive function, language, and visuospatial ability. Here, we provide a summary of each measure used as outcomes in the present study.

The California Verbal Learning Test (CVLT) [23] assesses episodic verbal learning and memory. There are 5 learning trials of 16 shopping items, presented orally, with four items from each of four semantic categories. The sum of the five trials provides a measure of new learning and immediate free recall. In addition, short- and long-delayed free recall, short- and long-delayed cued recall, and recognition memory are assessed. The two measures used were: total number of items recalled across the five immediate free recall trials (CVLT-IMM), and long-delay free recall (CVLT-LD), with maximum scores of 80, and 16, respectively.

The Benton Visual Retention Test (BVRT) [24] measures visual constructional skill and short-term figural memory. Participants study 10 line-drawings, including one to three geometric figures, for 10 seconds each, and then immediately reproduce them from memory using pencil and paper. The designs become more difficult over the 10 trials. The measure used was the total number of errors.

The Trail-Making Test Parts A (TMT-A) and B (TMT-B) [25] assess attention, concentration, visuomotor scanning, perceptuomotor speed, working memory, and set-shifting. TMT-A involves drawing a line to connect randomly arranged numbers from 1 to 25 in sequential order. TMT-B involves connecting randomly arranged numbers and letters in alternating sequence (e.g., 1-A-2-B …). Time to completion for each test (in seconds) was used in the present study.

Letter [26] and Category [27] Fluency are measures of fluent language production and executive function. Participants were given 60 seconds to generate as many words as possible beginning with specific letters and specific categories. The mean numbers of correct words generated in 60 seconds, across the three trials each for letter and category fluency, were the measures of interest.

The Boston Naming Test (BNT) [28] is a measure of object recognition and semantic retrieval. Participants identify and name a series of line drawings of objects, beginning with common objects and ending with infrequent objects. The measure used was the number of words out of 60 correctly named without cues.

WAIS-R Similarities measures verbal concept formation and abstract reasoning. [29] Participants are asked how 14 pairs of two items are similar, starting with concrete items and becoming increasingly abstract. The measure of interest was the total score out of a maximum of 28.

A modified version [30] of the Educational Testing Service Card Rotations test was used to measure visuospatial ability. Participants were presented with a target figure and eight alternative figures in the same row. Subjects marked images that could be rotated in plane to match the target, but not those that were mirror image figures. The total number correct minus total number incorrect across the two parts (14 targets per part) was the measure of interest, with a maximum score of 224.

The Mini Mental State Examination (MMSE) [31] assesses mental status, including orientation to time and place, immediate and delayed recall, attention and calculation, and language. Total score out of 30 was the measure of interest.

### Statistical analyses

To find the number of change-points where the rate of longitudinal decline changes significantly and the timing of these change-points relative to AD diagnosis, a series of linear and nonlinear mixed models with increasing complexity were fit with each of the cognitive measures as the outcome and the time (in years) to diagnosis of AD as the main predictor. We started with a no change-point model and then tested a one-change-point model and finally a two change-points model. The two-change-point model function is given by

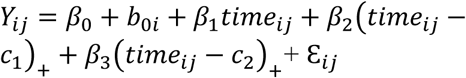

where (*x*)_+_ = *x, x* > 0 and (*x*)_+_ = 0, *x* < 0.

*c*_1_ is first change-point, *c*_2_ is second change-point. *b*_0*i*_ is a random effect that follows a normal distribution with mean 0 and standard deviation of σ.

All models included baseline age, sex (male vs. female), race (white vs. non-white), and years of education as covariates. Model selections were based on the likelihood ratio test. The best model fit tells us how many change-points, if any, there are for each cognitive measure.

The final model estimates were determined by a bootstrapping method so that the parameter estimates, and standard errors can be accurately captured, and more importantly, the timings of change-points from different cognitive measures can be statistically compared. We used 500 randomly resampled samples, with each sample having the same sample size as the original one for the bootstrapping. The models were fit using PROC NLMIXED in SAS 9.4 (Cary, NC).

## Results

Sample characteristics are shown in Table 1. The sample consisted of 165 participants with an AD diagnosis with a total of 988 visits. Average baseline age was 76.5 years (standard deviation, [SD]=7.4), the average follow-up interval was 8.3 years (SD=6.0, range=0-24.1), and average age at AD diagnosis was 86.5 years (SD=6.1). Eighty-three (50.3%) participants were female, and 85.5% of the sample were white. The average years of education was 16.7 years (SD=2.6, range=8-21).

**Table 1:**
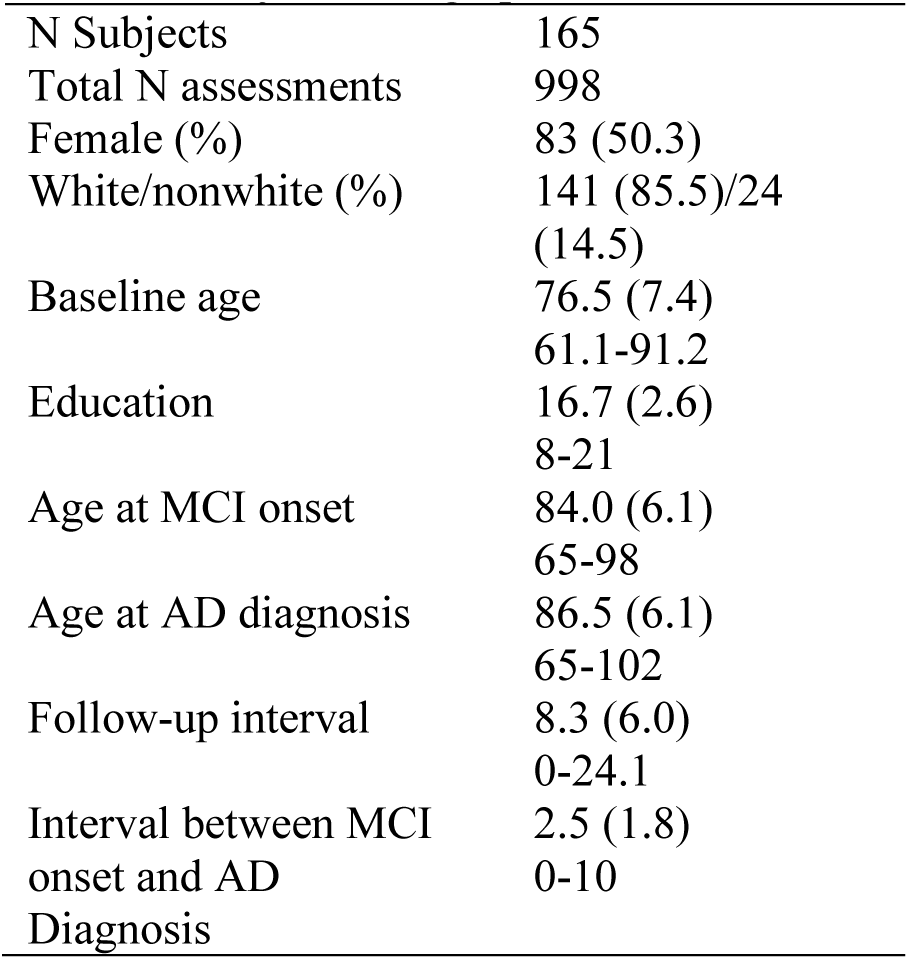
Subject demographics

### Change-point model comparisons for each cognitive measure

Table 2 contains the results of the model fit statistics (likelihood ratio test) comparing the fit of three models for each cognitive measure. Models with 1-change-point provided better fit compared with no change-point models for all cognitive measures. Models with 2-change-points provided a better fit for CVLT-IMM, CVLT-LD, Category Fluency, Letter Fluency, BNT, Similarities, Card Rotations, and the MMSE. However, for the BVRT, TMT-A, and TMT-B, the 2-changepoint model did not significantly improve the model fit, indicating a 1-change-point model was the best fitting model for these measures.

**Table 2:**
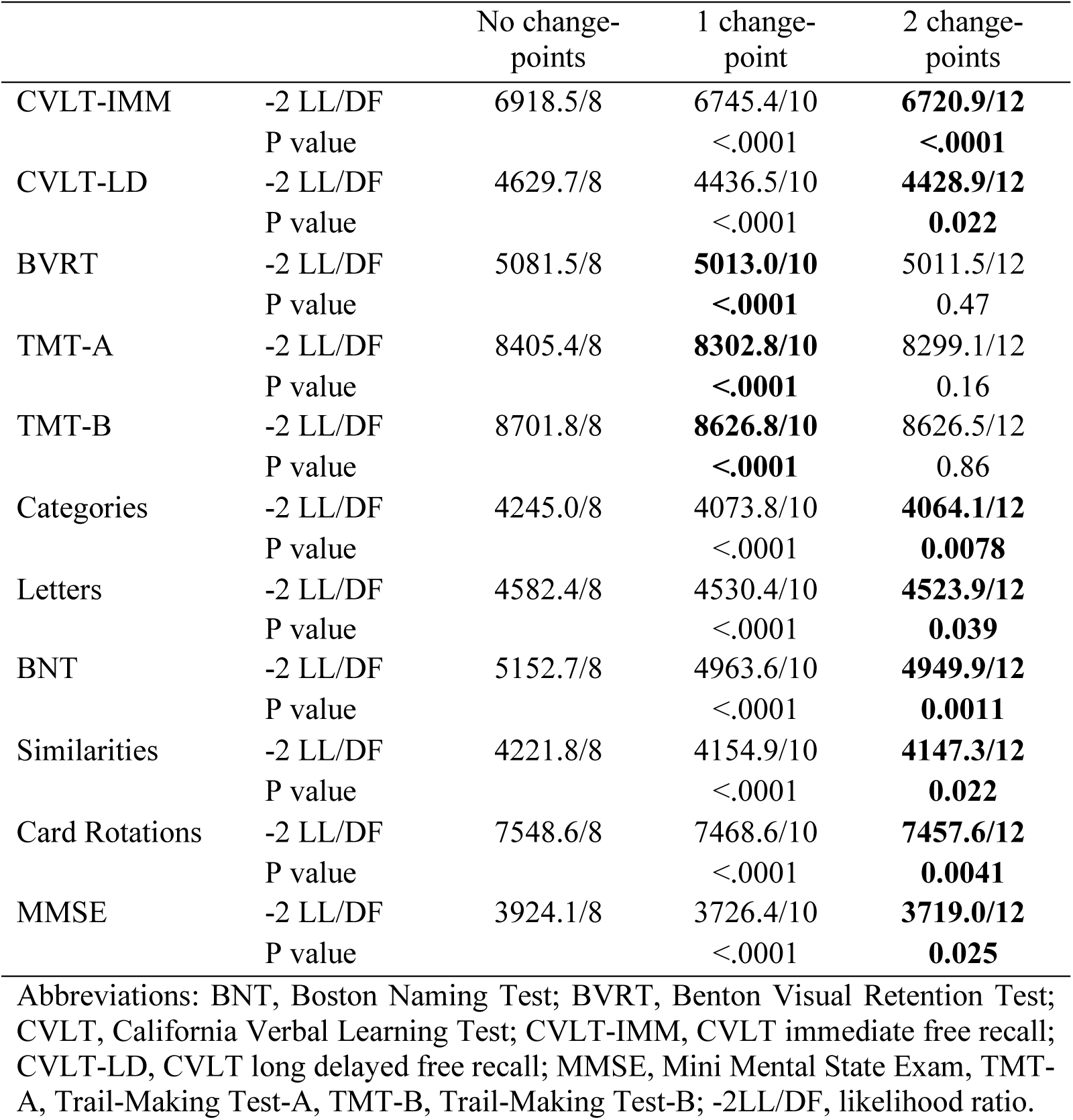
Fit statistics for models using no change-points, one change-point, or two change points. Model fit was assessed using the likelihood ratio test, with bold text indicating the current model provides significantly better fit compared to the one prior.

### Temporal position of change-points and subsequent rates of decline

Table 3 shows the results of parameter estimates from bootstrapping the best fitting model for each cognitive measure, including the estimated change-points, the rate of change at each segment of the trajectory, and corresponding standard errors (SE). Figure 1 shows the estimated trajectories superim-posed over the raw data for each cognitive measure.

**Table 3:** Estimated change-points in years from AD diagnosis and slopes in test units change per year. All estimates, standard errors, and p values are from bootstrapping.

| Estimate (S.E.) | Change-point 1 | Change-point 2 | Slope 1 | Slope 2 | Slope 3 |
| --- | --- | --- | --- | --- | --- |
| CVLT-IMM | -11.65 (0.80) | -2.80 (0.78) | 0.23 (0.28)<br>P = 0.41 | -1.44 (0.25)<br>P < .0001 | -3.74 (0.48)<br>P < .0001 |
| CVLT-LD | -7.58 (0.49) | -4.21 (0.33) | -0.080 (0.041)<br>P = 0.051 | -0.48 (0.13)<br>P = 0.0002 | -1.00 (0.091)<br>P < .0001 |
| BVRT * | -4.83 (0.81) | NA | 0.36 (0.034)<br>P < .0001 | 0.99 (0.12)<br>P < .0001 | NA |
| TMT-A* | -1.90 (0.68) | NA | 0.59 (0.14)<br>P < .0001 | 9.01 (3.12)<br>P = 0.0039 | NA |
| TMT-B* | -4.82 (0.73) | NA | 1.68 (0.42)<br>P < .0001 | 11.86 (2.21)<br>P < .0001 | NA |
| Categories | -9.89 (0.90) | -3.17 (0.41) | -0.13 (0.035)<br>P = 0.0002 | -0.34 (0.052)<br>P < .0001 | -0.97 (0.095)<br>P < .0001 |
| Letters | -10.03 (1.39) | -2.25 (0.91) | -0.010 (0.047)<br>P = 0.83 | -0.22 (0.058)<br>P = 0.00016 | -0.69 (0.20)<br>P = 0.00048 |
| BNT | -6.04 (0.74) | -1.51 (0.57) | -0.10 (0.036)<br>P = 0.0043 | -0.89 (0.17)<br>P < .0001 | -2.06 (0.57)<br>P = 0.00033 |
| Similarities | -10.65 (1.24) | -1.72 (0.53) | 0.038 (0.054)<br>P = 0.48 | -0.16 (0.043)<br>P = 0.00026 | -0.84 (0.23)<br>P = 0.00019 |
| Card Rotation | -15.48 (1.72) | -4.33 (1.18) | 1.66 (1.59)<br>P = 0.30 | -1.38 (0.35)<br>P < .0001 | -4.74 (1.08)<br>P < .0001 |
| MMSE | -9.13 (0.92) | -1.77 (0.44) | -0.038 (0.024)<br>P = 0.11 | -0.18 (0.033)<br>P < .0001 | -1.03 (0.15)<br>P < .0001 |

\* These measures only had a single change-point. Abbreviations: : BNT, Boston Naming Test; BVRT, Benton Visual Retention Test; CVLT, California Verbal Learning Test; CVLT-IMM, CVLT immediate free recall; CVLT-LD, CVLT long delayed free recall; MMSE, Mini Mental State Exam, TMT-A, Trail-Making Test-A; TMT-B, Trail-Making Test-B; S.E., standard error.

**Figure 1:**
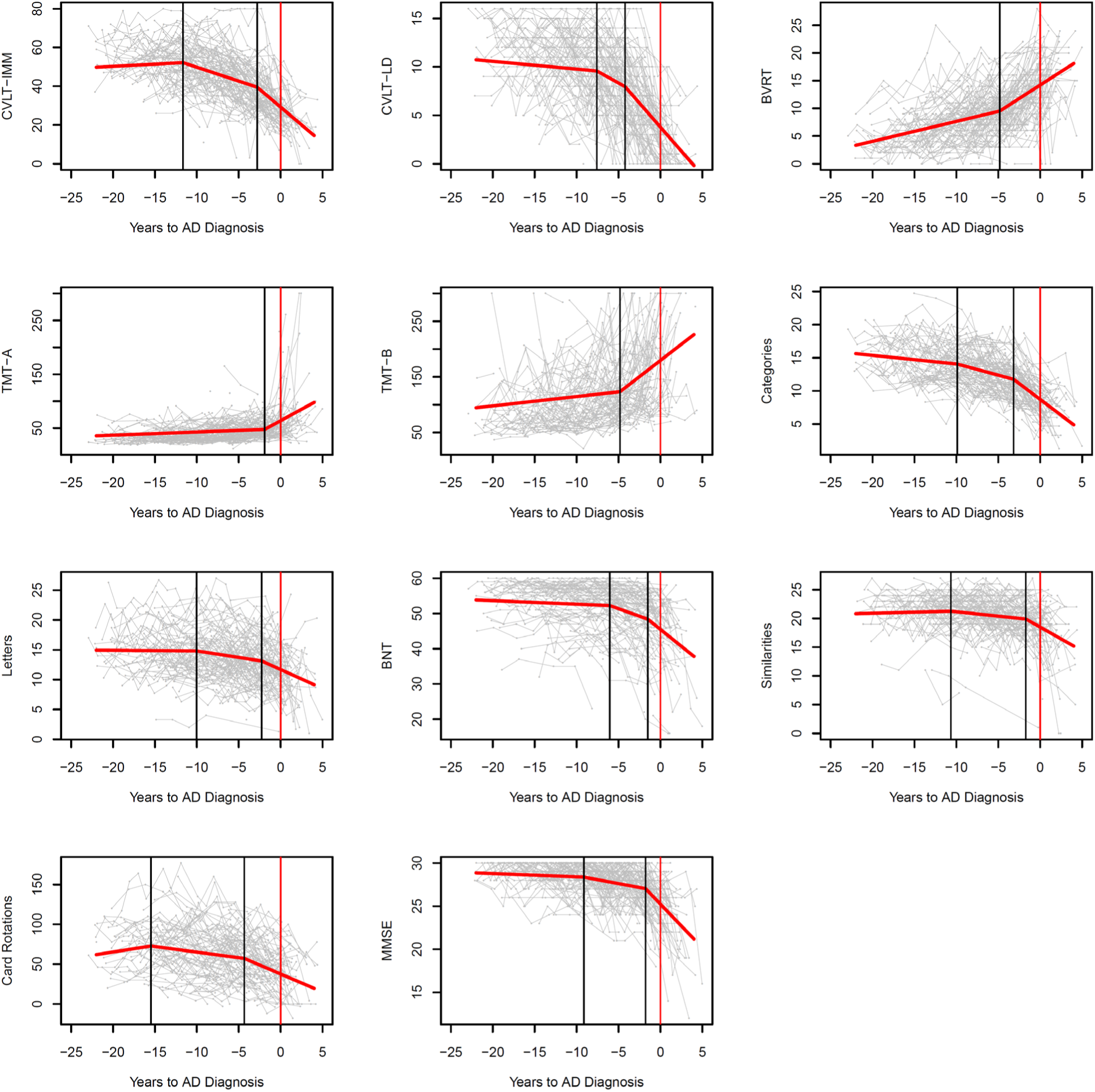
Line graphs showing the modelled population-level longitudinal trajectories from change-point models (in red) superimposed over spaghetti plots of the raw data (gray) for each cognitive measure. Vertical black lines indicate the change-points and the vertical red lines indicate timing of AD diagnosis. The X-axis represents years before AD diagnosis. Measurement units: CVLT-IMM, total correct out of 80; CVLT-LD, total correct out of 16; BVRT, total number of errors; TMT-A, seconds to complete; TMT-B, seconds to complete; Categories, mean number correct words; Letters, mean number correct words; BNT, correct out of 60; Similarities, total our of 28; Card Rotations, total out of 224; MMSE total out of 30.

### Learning and memory

#### CVLT-IMM

The estimated first change-point was 11.65 (SE=0.80) years before AD diagnosis, when the trajectory transitioned from non-significant increase in performance over time (0.23 items per year, SE=0.28) to significant moderate decline in performance (−1.44 items per year, SE=0.25). The second change-point was 2.80 (SE=0.78) years before AD diagnosis, when decline accelerated again (−3.74 items per year, SE=0.48).

#### CVLT-LD

The estimated first change-point was 7.58 (SE=0.49) years before AD diagnosis, when the trajectory transitioned from trending-significant minor decrease in performance over time (−0.080 items per year, SE=0.041) to significant moderate decline in performance (−0.48 items per year, SE=0.13). The second change-point was 4.21 (SE=0.34) years before AD diagnosis, when decline accelerated again (−1.00 items per year, SE=0.091).

#### BVRT

The estimated change-point was 4.83 (SE=0.81) years before AD diagnosis, when the trajectory transitioned from significant modest decline in performance over time (0.36 errors per year, SE=0.034) to significant accelerated decline in performance (0.99 errors per year, SE=0.12).

### Attention and executive function

#### TMT-A

The estimated change-point was 1.90 (SE=0.68) years before AD diagnosis, when the trajectory transitioned from significant modest decline in performance over time (0.59 seconds per year, SE=0.14) to significant accelerated decline in performance (9.01 seconds per year, SE=3.12).

#### TMT-B

The estimated change-point was 4.82 (SE=0.73) years before AD diagnosis, when the trajectory transitioned from significant modest decline in performance over time (1.68 seconds per year, SE=0.42) to significant accelerated decline in performance (11.86 seconds per year, SE=2.21).

### Verbal fluency

#### Category fluency

The estimated first change-point was 9.89 (SE=0.90) years before AD diagnosis, when the trajectory transitioned from significant minor decrease in performance over time (−0.13 words per year, SE=0.035) to significant moderate decline in performance (−0.34 words per year, SE = 0.052). The second change-point was 3.17 (SE=0.41) years before AD diagnosis, when decline accelerated again (−0.97 words per year, SE=0.095).

#### Letter fluency

The estimated first change-point was 10.03 (SE=1.39) years before AD diagnosis, when the trajectory transitioned from non-significant minor decrease in performance over time (−0.010 words per year, SE=0.047) to significant moderate decline in performance (−0.22 words per year, SE = 0.058). The second change-point was 2.25 (SE=0.91) years before AD diagnosis, when decline accelerated again (−0.69 words per year, SE=0.20).

### Object recognition and naming

#### BNT

The estimated first change-point was 6.04 (SE=0.74) years before AD diagnosis, when the trajectory transitioned from significant minor decrease in performance over time (−0.10 words per year, SE=0.036) to significant moderate decline in performance (−0.89 words per year, SE = 0.17). The second change-point was 1.51 (SE=0.57) years before AD diagnosis, when decline accelerated again (−2.06 words per year, SE=0.57).

### Abstract reasoning

#### Similarities

The estimated first change-point was 10.65 (SE=1.24) years before AD diagnosis, when the trajectory transitioned from non-significant increase in performance over time (0.038 points per year, SE=0.054) to significant moderate decline in performance (−0.16 points per year, SE=0.043). The second change-point was 1.72 (SE=0.35) years before AD diagnosis, when decline accelerated again (−0.84 points per year, SE=0.23).

### Visuospatial ability

#### Card Rotations

The estimated first change-point was 15.48 (SE=1.72) years before AD diagnosis, when the trajectory transitioned from non-significant increase in performance over time (1.66 points per year, SE=1.59) to significant moderate decline in performance (−1.38 points per year, SE=0.35). The second change-point was 4.33 (SE=1.18) years before AD diagnosis, when decline accelerated again (−4.74 points per year, SE=1.08).

### Global Cognitive Performance

#### MMSE

The estimated first change-point was 9.13 (SE=0.92) years before AD diagnosis, when the trajectory transitioned from non-significant increase in performance over time (−0.038 units per year, SE=0.024) to significant moderate decline in performance (−0.18 units per year, SE=0.033). The second change-point was 1.77 (SE=0.47) years before AD diagnosis, when decline accelerated again (−1.03 units per year, SE=0.15).

### Comparing change-points across cognitive measures

Figure 2 provides a schematic overview of estimated change-points for each cognitive measure. Table 4 shows the results from using bootstrapping to compare the first change-points for each measure against all other measures to identify the earliest changing measures. The measure with the earliest change-point was Card Rotations, which was significantly earlier than all other measures. The next measure to show an early change-point was CVLT-IMM, which was significantly earlier than CVLT-LD, BVRT, TMT-A, TMT-B, BNT, and MMSE but not significantly earlier than measures of verbal fluency, or Similarities. The measure to show the latest change-point in relation to AD diagnosis was TMT-A. Supplemental Table 1 shows the results from using bootstrapping to compare the second change-points for each measure. The second change-point for Card Rotations was not significantly earlier than the second change-points for CVLT measures.

**Table 4:** The first change-point for each measure was compared between all measures using bootstrapping. P-values are presented to show which measures were significantly different from each other.

|  | CVLT-IMM | CVLT-LD | BVRT | TMT-A | TMT-B | Categories | Letters | BNT | Similarities | Card<br>Rotation | MMSE |
| --- | --- | --- | --- | --- | --- | --- | --- | --- | --- | --- | --- |
| CVLT-IMM | 1 | <.0001 | <.0001 | <.0001 | <.0001 | 0.13 | 0.31 | <.0001 | 0.46 | 0.045 | 0.031 |
| CVLT-LD |  | 1 | 0.0029 | <.0001 | 0.0022 | 0.031 | 0.10 | 0.087 | 0.025 | <.0001 | 0.13 |
| BVRT |  |  | 1 | 0.0039 | 0.99 | <.0001 | 0.0018 | 0.26 | 0.00012 | <.0001 | 0.00069 |
| TMT-A |  |  |  | 1 | 0.00063 | <.0001 | <.0001 | <.0001 | <.0001 | <.0001 | <.0001 |
| TMT-B |  |  |  |  | 1 | <.0001 | 0.00057 | 0.27 | <.0001 | <.0001 | 0.00034 |
| Categories |  |  |  |  |  | 1 | 0.93 | 0.0014 | 0.64 | 0.0041 | 0.56 |
| Letters |  |  |  |  |  |  | 1 | 0.019 | 0.73 | 0.010 | 0.56 |
| BNT |  |  |  |  |  |  |  | 1 | 0.0017 | <.0001 | 0.013 |
| Similarities |  |  |  |  |  |  |  |  | 1 | 0.023 | 0.33 |
| Card Rotation |  |  |  |  |  |  |  |  |  | 1 | 0.0015 |
| MMSE |  |  |  |  |  |  |  |  |  |  | 1 |

Abbreviations: BNT, Boston Naming Test; BVRT, Benton Visual Retention Test; CVLT, California Verbal Learning Test; CVLT-IMM, CVLT immediate free recall; CVLT-LD, CVLT long delayed free recall; MMSE, Mini Mental State Exam, TMT-A, Trail-Making Test-A; TMT-B, Trail-Making Test-B.

**Figure 2:**
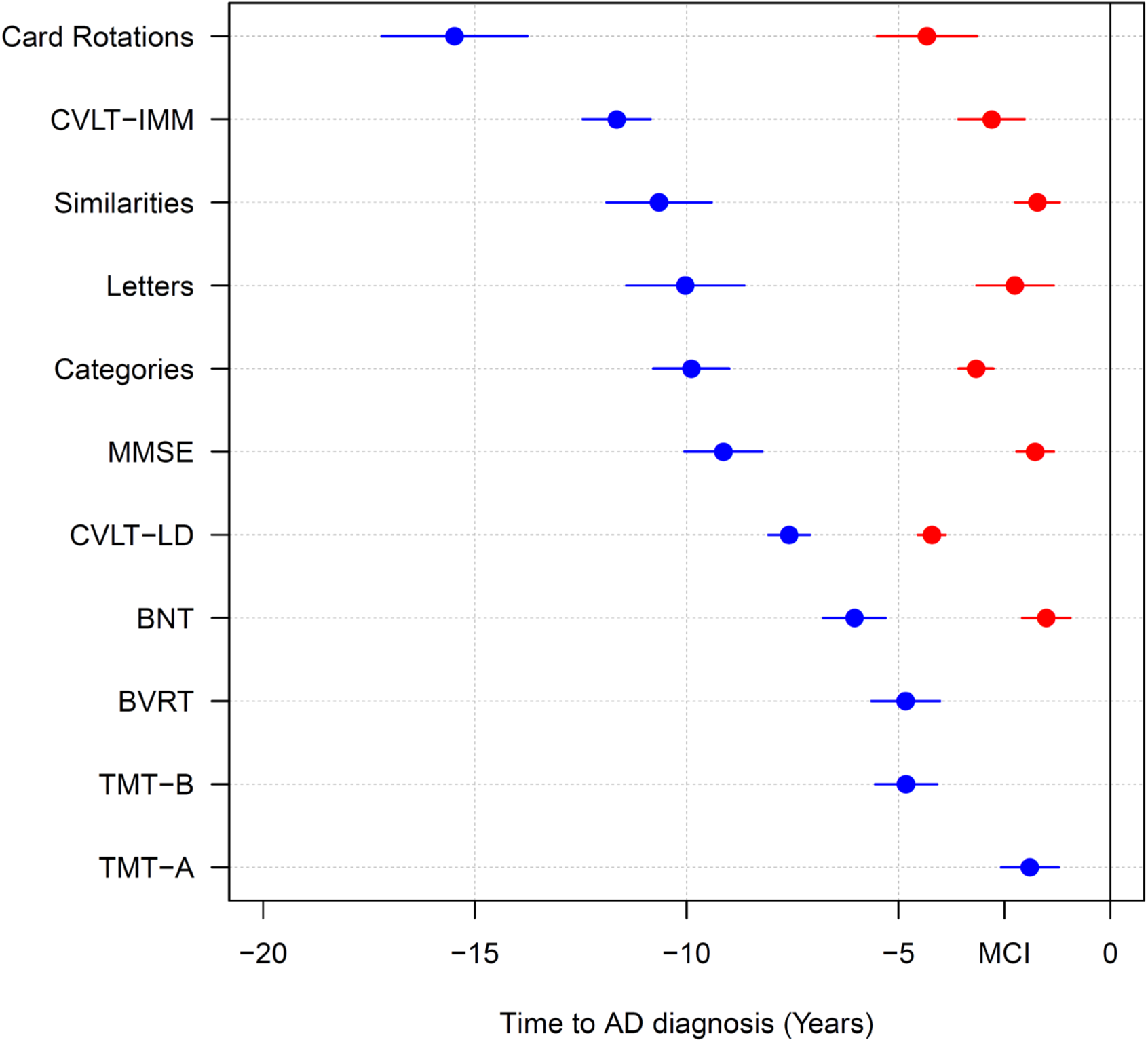
Dot plot comparing the estimated change points relative to AD diagnosis across cognitive measures. Cognitive measures are presented in order of first change-points. Red dots represent first change-points and blue points represent second change-points. Extended lines show standard errors. MCI indicates the average time of mild cognitive impairment symptom onset before AD diagnosis.

## Discussion

In a sample of participants with consensus diagnoses of clinical AD, we used extensive longitudinal cognitive data to examine the temporal sequence of stages of decline in 11 cognitive measures. Change-points identifying steeper rates of cognitive decline ranged from 15.5 years before AD diagnosis for the Card Rotations test to 1.9 years before AD diagnosis for TMT-A. While episodic memory assessed by CVLT measures was not the domain to show the earliest changes in rates of decline, changes were still detected up to 11.7 years before AD diagnosis. Using change-point analyses in this way can reveal the temporal ordering of domain-specific accelerated decline in preclinical AD.

The change-point for Card Rotations (15.5 years before AD diagnosis) was significantly earlier than change-points for all other cognitive measures, including CVLT measures of episodic memory. This extends previous findings from a systematic review [14] in which it was casually observed that visuospatial ability shows the earliest acceleration of cognitive decline prior to AD.

The underlying mechanisms that may lead to early accelerated decline in visuospatial ability in preclinical AD may be understood from the roles of the precuneus and other parietal regions in visuospatial tasks that involve spatial manipulation (as is the case with the Card Rotations Task in BLSA). [32] The precuneus is also part of a large network that includes medial temporal lobe and frontal lobe regions that support spatial navigation. [33] The precuneus is one of the earliest brain regions to show accumulation of Aβ in preclinical AD [34, 35] and deficits in spatial navigation are one of the earliest impairments leading to loss of independence. Taken together, the functional importance of the precuneus in visuospatial processing and its susceptibility to early AD pathology support our finding that visuospatial ability would be affected early in preclinical AD.

CVLT-IMM showed the second earliest change-point at 11.7 years before diagnosis. This change was significantly earlier than CVLT-LD, which had a change-point at 7.6 years before AD diagnosis. The difference between change-points for CVLT-IMM and CVLT-LD is consistent with previous studies that reported faster rates of verbal learning compared to delayed free recall declines at earlier stages of disease progression, [36, 37] and confirms the importance of early learning deficits in detecting individuals at risk of developing AD. [38] However, change-points for CVLT-IMM were not significantly earlier than those for measures of verbal fluency, the Similarities Test, or the MMSE, suggesting that some aspects of executive function, i.e., verbal concept formation and abstract reasoning, as well as aspects of mental status may exhibit changes in the rates of decline as early as some memory-based learning tasks. These results contrast with earlier reports using BLSA data indicating that memory is affected earlier than executive function. [13] One possible explanation for the different pattern of results in the present analysis is the larger number of participants with longer follow-up compared to previous reports.

Every cognitive measure examined showed at least one change-point, with the majority of measures exhibiting two change-points. In measures with two change points, the first change-point was always more than five years before diagnosis while the second change-point was less than five years before diagnosis and was followed by even faster rates of decline than the first change-point. The second change-points appear to represent the transition from preclinical AD to MCI. The mean time between symptom onset of MCI and AD diagnosis in our sample was 2.5 (SD=1.8) years. Accelerated cognitive declines in the years immediately prior to symptom onset and AD diagnosis are consistent with other reports that MCI participants show raster rates of decline compared to healthy controls for a range of cognitive domains including memory, executive function, attention and verbal fluency. [39]

The temporal ordering of the first and second change-points across cognitive measures were similar. However, the second change-point for Card Rotations was not significantly earlier than the second change-points for measures of memory or fluency, suggesting that there is little difference in the sensitivity of these measures as the time to diagnosis becomes shorter. As noted by Grober et al. [13] the temporal unfolding of cognitive decline identified by change-point studies implies that the predictive utility of different measures would be expected to vary by time from AD diagnosis. As such, the temporal ordering of change-points in the present study would suggest that measures of visuospatial ability and memory may serve as predictive tools for the development of AD as much as 15 years before diagnosis with other measures becoming more relevant closer to diagnosis. However, some measures of processing speed, i.e., TMT-A, may only have predictive utility less than five years before diagnosis, during a period when MCI may already be detectible. This interpretation is supported by previous reports of the predictive power of different cognitive measures. [40]

A limitation of this study is that BLSA participants are a highly educated group, which may limit generalizability. Additionally, the focus of this study was on defining and comparing population-level change-points and as such we did not examine individual differences in the timing of change-points which requires longer follow-up at the individual level. Future work is needed to assess individual differences in change-points. However, the strengths of this study are the comprehensive cognitive battery, frequent visit schedule (subjects were tested annually or biannually in this sample) and consensus-based determination of symptom onset and AD diagnosis. In addition, the use of the bootstrapping analysis not only allows us to capture parameter estimates and standard errors more accurately, but enables the statistical comparison of change-points among different cognitive measures, greatly extending the work of previous studies. [14]

In summary, we found that the cognitive measure to show the earliest change in rates of decline in preclinical AD was visuospatial ability rather than episodic memory. Using change-point analyses with bootstrapping can reveal the temporal patterns of accelerated cognitive decline in preclinical AD and may help guide the development of tools for participant screening in clinical trials.

## Data Availability

Data from the BLSA are available on request from the BLSA website (blsa.nih.gov). All requests are reviewed by the BLSA Data Sharing Proposal Review Committee and are also subject to approval from the NIH institutional review board.

## Acknowledgements

This research was supported entirely by the Intramural Research Program of the National Institutes of Health, National Institute on Aging. We thank the staff of the BLSA and LBN cognitive testing group for their assistance and the BLSA participants for their dedication to this study.

